# SEAD: an augmented reference panel with 22,134 haplotypes boosts the rare variants imputation and GWAS analysis in Asian population

**DOI:** 10.1101/2023.12.23.23300480

**Authors:** Meng-Yuan Yang, Jia-Dong Zhong, Xin Li, Geng Tian, Wei-Yang Bai, Yi-Hu Fang, Mo-Chang Qiu, Cheng-Da Yuan, Chun-Fu Yu, Nan Li, Ji-Jian Yang, Yu-Heng Liu, Shi-Hui Yu, Wei-Wei Zhao, Jun-Quan Liu, Yi Sun, Pei-Kuan Cong, Saber Khederzadeh, Pian-Pian Zhao, Yu Qian, Peng-Lin Guan, Jia-Xuan Gu, Si-Rui Gai, Xiang-Jiao Yi, Jian-Guo Tao, Xiang Chen, Mao-Mao Miao, Lan-Xin Lei, Lin Xu, Shu-Yang Xie, Jin-Chen Li, Ji-Feng Guo, David Karasik, Liu Yang, Bei-Sha Tang, Fei Huang, Hou-Feng Zheng

## Abstract

Here, we present the **S**outh and **E**ast **A**sian reference **D**atabase (**SEAD**) panel (https://imputationserver.westlake.edu.cn/), which comprises whole genome sequencing data from 11,067 individuals across 17 countries in Asia. The SEAD panel, which excludes singleton variants, consists of 22,134 haplotypes and 88,294,957 variants. The SEAD panel demonstrated higher accuracy compared to 1kGP, TOPMed and ChinaMAP in South Asian population. And as the proportion of South Asian ancestry increased, the proportion of low-frequency and rare well-imputed variants imputed using the SEAD panel progressively increased, whereas those imputed with TOPMed panel significantly decreased. Additionally, when imputing the East Asian population, the SEAD showed comparable concordance in imputation with ChinaMAP panel, while the TOPMed panel was inferior. Finally, we applied the augmented SEAD panel to conduct a discovery and replication genome-wide association study (GWAS) for hip and femoral neck (FN) bone mineral density (BMD) traits within the 5,369 Westlake BioBank for Chinese (WBBC) genotyped samples. The single-variant test suggests that rare variants near *SNTG1* gene are associated with hip BMD (rs60103302, MAF=0.0092, *P*=1.67×10^−7^). The variant-set analysis also suggests the association of this gene (*P*_slide_window_=9.08×10^−9^, *P*_gene_centric_=5.27×10^−8^). The gene and variants achieved a suggestive level for FN BMD. This gene was not reported previously and can only be detected by using Asian reference panel. The preliminary experiment in-vitro demonstrated that the identified rare variant could upregulate the *SNTG1* expression, which in turn inhibits the proliferation and differentiation of preosteoblast.

## Introduction

Genotype imputation is an effective way to save the cost of sequencing and has become a key step in genome-wide association studies (GWAS). It can increase the chance of identifying likely causal variants (1), facilitate meta-analysis (2, 3), and aid in finding pleiotropic effects of risk variants (4, 5). Factors such as the haplotype size, ancestry diversity and sequencing depth can affect the imputation accuracy (6). Generally, a diverse reference panel can improve accuracy in genetically diverse populations (7), while an ancestry-specific reference panel can benefit the corresponding population (8, 9). In our pervious study, we found that the imputation accuracy for European population benefit from an increasing in haplotype size and population diversity of the reference panel, while the accuracy for the Han Chinese population reached its peak with a fraction of an extra diverse sample (8-21%) in the reference panel (10). Given that imputation accuracy directly impacts the credibility of subsequent analysis, it is crucial to select an appropriate reference panel before imputation.

The 1000 Genomes Project (1kGP) is one of the most renowned and widely used reference panel for genotype imputation, comprising 2,504 individuals from 26 populations (11). The latest update to 1kGP phase 3 sequenced 3,202 samples (including an additional 698 samples related to the previous participants) and identified 70,594,286 single nucleotide polymorphisms (SNPs) with 30× depth. As many rare and low-frequency variants associated with diseases tend to be population-specific (11), an increasing number of human genome projects are focusing on specific populations to provide population-specific reference panels. However, most of the whole-genome sequencing (WGS) efforts were carried out in European-descent individuals, such as the GoNL project (the Genome of the Netherlands project, n=769) (12), UK10K project (∼4,000 WGS and ∼6,000 whole exome sequencing samples in UK) (13, 14) and the TOPMed program (Trans-Omics for Precision Medicine, n=∼138,000) (15). These efforts have enabled the provision of a large-scale combined reference panel for the European population, such as the HRC panel (Haplotype Reference Consortium, n=32,470) (16). The update of analytical methods and the emergence of various imputation reference panels have improved the imputation quality for European population. However, one of the complicated legacies of human genome project after 20 years’ development is that lacking of diversity might hinder the promise of genome science (17). As the largest continent, Asia accounts for 59.5 % of the worldwide population (https://www.worldometers.info/world-population/). Fortunately, the Asian ancestry-dominant populations have been recently sequenced and analyzed to understand the genetic basis of the Asian, such as the Japanese (18), Korean (19), Singaporean (20), Indian (21) and Chinese (22–25). Among these efforts, the Westlake BioBank for Chinese (WBBC) project (24, 26) was initiated in 2017 by our team. Up to now, we have 4480 Chinese samples whole-genome sequenced (WBBC-seq) and 6080 samples whole-genome genotyped (WBBC-chip), covering 29 out of 34 administrative divisions of China (24, 27).

Rare variants, which may have low level of pairwise linkage disequilibrium with common variants, can potentially result in significant functional consequences (28). Next-generation sequencing, with adequate coverage, enables the accurate detection of rare variants (29). Rare variants are harder to impute than common variants because rare variants only appear a few times in the reference panel. Further, the evaluation of rare variant imputation in Asian populations is hindered by the limited sample size of currently published studies, which typically comprise only a few thousand individuals.

Recent evaluations of global populations by TOPMed revealed that even with a sample size of over 130,000 individuals, including 14,000 Asian samples, the imputation accuracy for some Asian populations remains insufficient (30). In this study, we integrated the WGS data from multiple sources, including Singapore SG10K pilot project (13.7×, 4563 samples), GenomeAsia pilot project (36×, 1031 samples), WBBC pilot project (13.9×, 4480 samples), and the high-coverage 1kGP-Asian phase 3 (30×, 993 East and South Asian). These data were combined to create the **S**outh and **E**ast **A**sian reference **D**atabase (**SEAD**) panel, encompassing a total of 11,067 individuals, making it a large-scale reference panel for Asian populations. We then focused on evaluating the imputation quality of this panel to impute rare variants in Asian populations. Finally, we applied this augmented panel to the WBBC-chip data to explore its role in imputing possible likely causal rare variants for bone-related traits.

## Methods

### Variants calling and quality control for the Asian panels

We collected Asian whole genome sequencing data from the Westlake Biobank for Chinese (WBBC) pilot project (24), the newly released high-coverage 1000 Genomes project (1kGP) (31), SG10K pilot project (20) and the GenomeAsia pilot project (GAsP) (21) (Figure 1 Step 1).

The WBBC dataset, which were generated by our own, contains both whole genome sequencing (WBBC-seq) data and high-density Asian Screening Array (ASA) chip data (WBBC-chip) data (24). The WBBC-seq dataset included 4,480 Chinese samples with an average sequencing depth of 13.9×. The CRAM files of WBBC-seq were used in constructing the SEAD panel and had been stored locally. The CRAM files for the 993 EAS and SAS ancestry samples (1kGP-Asian) were downloaded here (ftp://ftp.sra.ebi.ac.uk/vol1/run/ERR323/). We performed variant calling on the 4480 samples of WBBC-seq and 993 samples of 1kGP-Asian using DeepVariants (32). We used bcftools filter to segment each chromosome of each sample into 2.5Mb subsets, followed by joint calling of the same regions across all samples using GLnexus (33). This resulted in genotype files for 5,473 WBKG (WBBC-seq and 1kGP-Asian) samples. Following our previous methods (24), we conducted quality control on the obtained genotype files. We used bcftools filter to remove SNVs and INDELs close to INDELs with SnpGap 3 and IndelGap 5, and filtered out INDELs longer than 50 bp. Individual genotypes with a genotype quality score (GQ) < 20 were set to missing. Missing and low-confidence genotypes were refined using BEAGLE 5.173 (34). The chromosomes were divided into chunks of 1Mb with 0.1Mb overlapping. Lastly, we excluded variants with a Hardy-Weinberg equilibrium (HWE) *p*-value < 1×10^−6^ using VCFtools (v 0.1.13) (35). The high-coverage 1kGP dataset (2504 samples) with an average depth of 30×, based on the GRCh38 assembly, were downloaded from the https://www.internationalgenome.org/data-portal/data-collection/30x-grch38. This dataset, not 1kGP-Asian, was used in the comparison of imputation performance of different reference panels (TOPMed, ChinaMAP, 1kGP and SEAD).

The SG10K dataset comprises 4,810 whole-genome sequencings (WGS) of Singaporean Chinese, Malays, and Indians, with an average depth of 13.7×. Since the SG10K were based on Genome Reference Consortium Human build 37 (GRCh37), we employed Picard LiftoverVcf (http://broadinstitute.github.io/picard/) to update the genome assembly version from GRCh37 to GRCh38. We merged the 22 autosomes and set the following parameters: java -Xmx100g - XX:ParallelGCThreads=8 -jar picard.jar LiftoverVcf I=inpute.vcf.gz O=SG10K_all.hg38.vcf.gz CHAIN=hg19ToHg38.over.chain.gz REJECT= rejected_variants.vcf R=Homo_sapiens_assembly38.fasta. The GAsP dataset includes 1,163 WGS samples with an average depth of 36×, mainly covering Indians, Korean, Pakistanis, etc. The downloaded VCF files in the GAsP database are categorized into three types based on variant types: INDELs, multiallelic, and SNVs. Initially, each VCF file underwent a coordinate transformation from GRCh37 to GRCh38 using Picard LiftoverVcf. Subsequently, genotype phasing was performed using Shapeit v2 with parameters including windows of size 0.5, 200 states, and an effective size of 14,269, followed by normalization with BCFtools (36). The multiallelic variants were split into biallelic forms and positions marked with asterisks were removed. Finally, the three different types of VCF files were merged using BCFtools concat.

We identified relative pairs using KING (37) and excluded sample pairs with estimated kinship coefficients restricted to the range of 0.177-0.354. Ultimately, we retained 4480 samples from WBBC-seq, 993 samples from 1kGP-Asian, 4563 samples from SG10K and 1031 samples from GAsP (Figure 1 Step 1, and Supplementary Table 1).

### Batch effect evaluated by principal component analysis

Given the differences in sequencing, batch processing, and quality control across the four datasets (WBBC-seq, 1kGP-Asian, SG10K and GAsP), we evaluated the batch effect by principal component analysis (PCA). We initially conducted a PCA on the WBKG dataset obtained through joint calling mentioned above. Subsequently, we merged the WBKG dataset with the SG10K dataset. We converted the processed VCF files into PLINK formats separately, following the merging guidelines available at https://github.com/baharian/merge_PLINK. The pipeline initially examined each dataset individually, inspecting each chromosome for SNPs identified by multiple or different names (e.g., rsID) at the same position, and removed any such occurrences. Subsequently, variants with missing calls were identified and removed from each dataset. The intersection of SNPs presenting in all datasets was determined, and SNPs not in this intersection were excluded. Next, the datasets were cross-examined for SNPs that might pose issues due to strand misassignment, reference/alternate allele misassignment, ambiguous strand or reference/alternate allele misassignment, and non-bi-allelic variants. Appropriate actions, such as flipping or removing SNPs, were taken for each category. After merging these datasets, an in-depth LD-based strand assignment cross-check was conducted on the merged data to identify and correct potential strand flips. We performed another PCA on the combined dataset (WBKG and SG10K). Finally, following the same methodology, we merged this dataset with the GAsP dataset and conducted another PCA analysis. The PLINK2.0 was used to compute the 20 principal components (PCs) with the parameters --pca approx 20 --maf 0.01 on the merged data. To further validate the PCA results, we extracted SNPs from the GAsP dataset with GQ > 40 and DP > 20 and computed the PCs using the same parameters.

### Quality control of the WBBC-chip data

Besides the whole genome sequencing data, the WBBC study also genotyped 6,080 individuals with the high-density Illumina Asian Screening Array (ASA, based on GRCh37), resulting in the identification of a total of 659,184 SNPs (24, 26). Of which, 184 samples underwent both whole-genome sequencing and array genotyping.

As part of quality control, we employed GCTA (38) to calculate the pairwise genetic relationship matrix using common variants and remove samples with a coefficient > 0.025 (Supplementary Table 3). We then excluded samples with missing call rates ≥ 5% and excluded SNPs with missing call rates ≥ 5%, MAF<1% and Hardy-Weinberg equilibrium at *P* < 1 × 10^−6^ by using PLINK1.9 (39). The genotype assembly version of the WBBC-chip data was updated from GRCh37 to GRCh38. Finally, we retained a total of 470,242 variants and 5,679 samples in the WBBC array dataset. The data were phased using SHAPEIT v2 with a window size of 0.5, 200 states, and an effective size of 14,269.

### Evaluation of imputation performance in UK biobank Asian dataset

To evaluate the imputation performance of SEAD panel in South Asian (SAS) population, we used Rye software (40) to infer the genetic ancestry composition of each sample in UK biobank, and selected 4,758 samples with 50%-70% SAS ancestry composition, 2,079 samples with 70%-90% SAS composition, and 2,205 samples with more than 90% SAS composition (Figure 1 Step 2). In brief, the UK Biobank data was directly downloaded upon request (Application 41376) by our team (41–47), and the 1kGP phase 3 data including 2,504 samples from five ancestral groups, Africa (AFR), East Asia (EAS), Europe (EUR), South Asia (SAS) and the Americas (AMR) (11), served as the ancestry reference populations. Rye infers genetic ancestry composition based on principal component (PC) analysis of samples from ancestral reference populations, and compares them to the person being tested. To obtain PCs of each individual, we merged the UK Biobank data with the 1kGP phase 3 data using PLINK2 and extracted 565,631 slightly LD-pruned HapMap3 variants to calculate the first 20 principal components (PCs). The LD-pruning parameters used in PLINK2 were: window size=1000 kb, step size=100, r^²^=0.9 and MAF≥0.01 (48, 49).

For the selected SAS ancestry samples in UK biobank, we extracted Asian Screening Array variants (659,184 variants) and converted the genotype coordinates to GRCh38 reference to produce the pseudo ASA arrays. We applied parameters --geno 0.05, --hwe 1e-6, --maf 0.01, -- make-bed, and --mind 0.05 to do the quality control in PLINK2. Finally, the samples with 50%-70% South Asian (SAS) ancestry retained 610,279 variants, those with 70%-90% SAS ancestry retained 583,566 variants, and those with >90% SAS ancestry retained 576,772 variants. We employed SHAPEIT v2(50) for phasing and conducted imputation locally using the SEAD and 1kGP panels respectively. For the imputation with TOPMed (https://imputation.biodatacatalyst.nhlbi.nih.gov/) and ChinaMAP (http://www.mbiobank.com/imputation/help/) panels, we submitted the phased genotypes to the online websites for processing. All these analyses were performed on chromosome 2.

### Evaluation of genotype concordance in HGDP Asian dataset

The 929 genomes from 55 diverse human populations from Human Genome Diversity Project (HGDP) (51) dataset was utilized to estimate the genotype concordance of imputation across Asian populations (Figure 1 Step 2). The HGDP dataset includes 104 samples from African populations, 61 samples from American populations, 197 samples from Central and South Asia populations, 223 samples from East Asia populations, 155 samples from European populations, 161 samples from Middle East populations, and 28 samples from Oceania populations (Supplementary Table 2). We obtained the HGDP data from the following source: ftp://ngs.sanger.ac.uk/production/hgdp. The phased autosome VCF file was split into 22 chromosomes, which were considered as the true set. To compare the imputation concordance of the reference panels (TOPMed, ChinaMAP, 1kGP, and SEAD), We extracted Asian Screening Array sites (659,184 variants) in Central and South Asia populations (197 samples) and East Asia populations (223 samples) as Asian array for comparison purpose. Before imputation, variants in the pseudo arrays with MAF less than 1% and samples with calling rate below 95% were excluded. We calculated the non-reference heterozygote concordance rate, non-reference homozygote concordance rate, precision and non-reference sensitivity between true sequencing data) and imputed genotypes pseudo arrays for each individual (Supplementary Figure 1), as what we did before (24). All these analyses were performed on chromosome 2.

### Evaluation in Chinese population

The procedure to assess the imputation performance in the Chinese population using the reference panels (TOPMed, ChinaMAP, 1kGP and SEAD) was similar to the evaluation in HGDP dataset (Figure 1 Step 2). As we have thousands of WBBC-chip samples, we could assess the imputation performance for low-frequency and rare variants. Genotype imputation was performed using Minimac4 (52, 53), with a chunk length of 20 Mb and a chunk overlap of 4 Mb. We used R-square as the estimated value, defined as the squared correlation between imputed genotypes and observed genotypes, produced by Minimac4. Sites with an imputed r-square (Rsq) value greater than 0.8 were considered well-imputed variants. We grouped the variants into seven Minor Allele Frequency (MAF) bins: <0.1%, 0.1%-0.3%, 0.3%-0.5%, 0.5%-0.7%, 0.7%-1%, 1%-2% and 2%-5%. We counted the number of well-imputed variants and calculated the average Rsq within each MAF bin. This analysis performed on chromosome 2.

In the WBBC-chip data, 179 samples underwent both whole-genome sequencing and array genotyping. We evaluated the imputation quality by comparing the concordance rate in the 179 samples, while the genotypes obtained from WGS served as the true set. Employing the methods from the previous section, we calculated the non-reference heterozygote concordance rate, and non-reference homozygote concordance rate, precision and non-reference sensitivity for each individual.

### Meta-imputation

We initially generated four independent non-singleton reference panels for the WBBC, 1kGP, SG10K, and GAsP datasets using minimac3 (Figure 1 Step 2). Subsequently, we imputed the 5679 WBBC-chip data with these four reference panels, utilizing minimac4 with the parameter set to -meta to produce the empiricalDose.vcf.gz file format for subsequent meta-analysis. Finally, we merged the imputation results from the four panels using MetaMinimac2 (https://github.com/yukt/MetaMinimac2)(54).

### Genome wide association study of BMD traits

As we mentioned above, the WBBC pilot study genotyped 6,080 individuals with ASA array (WBBC-chip), and a bunch of bone-related phenotypes were collected within WBBC (26). After quality control, a total of 470,242 variants and 5,679 samples were retained. Here, we took two correlated bone mineral density (BMD) traits (total hip and femoral neck BMD) as example, we removed individuals with missing phenotype data, and excluded outliers using the mean ± 4 standard deviations (n=5,369 left) (Figure 1 Step 3). We then grouped the ASA samples according to the assessment place of collection: 2,332 samples recruited from Jiangxi province (samples were mainly from Southern China) were taken as discovery cohort (WBBC-chip1), and 3,037 samples recruited from Shandong province (samples were mainly from Northern China) were taken as replication cohort (WBBC-chip2), and *vice versa*. The baseline statistics of the study samples were shown in Supplementary Table 4. Comprehensive GWAS analyses were conducted with the imputed genotypes from the augmented SEAD reference panel. We kept the imputed variants with Rsq >0.5 in the GWAS analysis. The imputation boosted the analyzed genetic variants from 470,242 to 19,235,129, with 3,195,758 variants with MAF between 0.1% and 1%. The BMD phenotypes (total hip and femoral neck BMD) in this study was analyzed as continuous outcomes, adjusting for ‘sex’, ‘age’, ‘BMI’, ‘geographical region’, and ‘10 principal components (PCs)’. The parameters applied in PLINK were ‘--geno’ of 0.05, ‘--mind’ of 0.05, ‘--hwe’ of 1■×■10^−6^ and ‘--maf’ of 0.001. Furthermore, we performed the meta-analysis of GWAS summary statistics in cohort 1 and cohort 2 using METAL software (55). Rare variants (0.001<MAF<0.01) were considered as candidates if they met the following criteria: a *P*-value less than 1e-04 in either cohort, a *P*-value less than 0.05 in the other cohort, and a *P*-value less than 1e-04 in the meta-analysis. Concurrently, we performed GWAS analysis on hip BMD using the WBBC-chip imputed with ChinaMAP, SG10K, WBBC-seq, TOPMed, 1kGP and GAsP panel. All processing steps were consistent with those used for the data imputed with SEAD panel.

### Variant-set analysis for association using annotation information

We performed the STAAR (variant-set test for association using annotation information) (56) framework to identify rare variants that would be associated with BMD traits (Hip and FN BMD) (Figure 1 Step 3). The STAAR pipeline facilitates rare variants variant-set analyses, including sliding window-based analysis and gene-centric analysis (56). In each test, we included variants with a minor allele frequency (MAF) of 0.1% to 1%. We employed fixed-size sliding window analysis, allowing for the systematic examination of distinct genomic regions by moving a 2kb window every 1kb across the genome, a total of 1,392,296 windows was divided. The gene-centric analysis method is the variant-set test for association using annotation information for a gene (STAAR-O), enhancing the power of rare variant association test by incorporating multiple variant functional annotations. For the gene-centric non-coding variants, aggregation was performed based on 7 categories: downstream, enhancer variants overlaid with Cap Analysis of Gene Expression (CAGE) sites, promoter CAGE, enhancer variants overlaid with DNase hypersensitivity (DHS), promoter DHS, upstream, and UTR2, and clustering regions with rare variations less than 2 were excluded. Both the sliding window and gene-centric methods were performed separately on WBBC-chip1 and WBBC-chip2.

### Cell culture

The HEK293T cells were cultured at 37 °C in a humidified atmosphere with 5% CO2 using DMEM basal media supplemented with 10% fetal bovine serum. The MC3T3-E1 subclone 14 cells were cultured with similar condition with ascorbate-free αMEM basal media supplemented with 10% fetal bovine serum. The media were refreshed every 3-4 days, and cells were passaged when they reached 90% confluence. To initiate differentiation, the media were replaced with a calcification-inducing medium containing 50 ng/L vitamin C (VC), 10 nM dexamethasone, and 10 mM beta-glycerophosphate.

### Luciferase Reporter assays

Dual-Luciferase reporter assays were conducted following previously described methods (57). In summary, the wild type luciferase plasmid was constructed by inserting the sequence of hg38 chr8: 49957722-49958204 into pGL6-TA firefly luciferase reporter vector (Beyotime). The risk luciferase plasmid was designed according to the *SNTG1* intronic SNP rs111829635 (hg38, chr8:49958116 C-T). The 293T or MC3T3-E1 cells were seeded in a 24-well plate for 16 hours and subsequently transfected with luciferase reporter plasmids along with the control Renilla plasmid (Beyotime), using lipofectamine 3000 (Thermo Fisher Scientific). After 48 hours of transfection, cells were collected and lysed using the lysis buffer provided in the Dual-Luciferase Reporter Assay System kit (Beyotime). Luciferase activity was analyzed in accordance with the guidelines outlined in the technique manual (Beyotime).

### *SNTG1* overexpression and qRT-PCR analysis

The cDNA of mouse *SNTG1* was amplified via polymerase chain reaction (PCR) and subsequently cloned into the pEF-GFP vector (a gift from Connie Cepko, Addgene plasmid # 11154 (58)), by replacing GFP to create vector pEF-SNTG1. To initiate the overexpression of *SNTG1*, MC3T3-E1 cells were plated in 6-well and 24-well plates at a density of 6,000 cells per square centimeter. Following an incubation period of 16-20 hours, transfection of pEF-SNTG1 was performed using Lipofectamine 3000 transfection reagent (Thermo Fisher Scientific). As a negative control, pEF-GFP was utilized. Additionally, a set of duplicate wells without any transfection was included and labeled as “WT”. After a further 4-6 hours, the cell culture medium was replaced with differentiation-induction medium.

For quantitative real-time polymerase chain reaction (qRT-PCR) analysis, cells were harvested 72 hours after transfection. Total RNA was extracted from the target cells using TRIZOL (Invitrogen) according to the manufacturer’s protocol. The isolated RNA was then reverse transcribed into complementary DNA (cDNA) using a reverse transcription kit (TransGen Biotech). For qRT-PCR analysis, a 2xSYBR Green Mix (TransGen Biotech) was utilized. The expression level of the target gene was normalized to that of GAPDH, which served as an endogenous control, enabling the comparison of samples. Cells intended for alkaline phosphatase (ALP) activity measurements were harvested after 6 days following transfection.

## Results

### Integration of the SEAD reference panel

First, we conducted SNP calling and joint calling to merge WBBC-seq (4480 samples) and 1kGP-Asian (993 samples) datasets by using DeepVariants and GLnexus. The number of variants identified by joint calling for WBBC-seq individuals ranged from 3,220,000 to 3,400,000 with a transition/transversion (ts/tv) ratio of 2.136-2.152. For the WBBC and 1kGP (WBKG) joint-calling, the variant count ranged from 3,000,000 to 3,250,000 with a ts/tv ratio of 2.142-2.162, indicating that the number and quality of variants were within reasonable limits (Supplementary figure 2). Additionally, we evaluated the sequencing quality by comparing the concordance rate in the 179 samples who had both WBBC-seq and WBBC-chip data, where taking the chip data as “true”. The majority of samples exhibited very high concordance rate around 0.99 for non-reference allele, heterozygote and homozygote genotypes, validating the use of these variants for further evaluation (Supplementary figure 3). After excluding singletons, the WBKG panel contained 5,473 samples and 43,542,610 variants.

Because of limited access to the SG10K and GAsP data, we applied reciprocal imputation approach with the similar protocol as the UK10K did (59): first, we merged the reference panel of SG10K (4,563 samples and 95,597,234 variants) and WBKG by imputing with each other, we then combined haplotypes from GAsP (1,031 samples and 63,925,145 variants) to the WBKG-SG10K panel. Finally, we obtained the **SEAD** (**S**outh and **E**ast **A**sian reference **D**atabase) panel by combining all the panels together (Figure 1 Step1).

Since the SEAD panel was derived from 4 different datasets, we evaluated the batch effect by principal component analysis (PCA). The results showed that the WBBC-seq matched very well with the 1kGP-EAS (Supplementary Figure 4A). The SG10K (comprising three populations: Indonesian, Chinese and Indian) and the GAsP (including a more diverse range of Asian populations), clustered with EAS and SAS, and populations in between (Supplementary Figure 4B and 4C). Further extraction of sites with genotype quality (GQ) greater than 40 and depth of coverage (DP) greater than 10 from the GAsP population produced almost identical results (Supplementary figure 4D), confirming the absence of batch effects in the datasets used for constructing the haplotype reference panel.

After removing the singletons, we finally got 88,294,957 variants and 22,134 haplotypes for SEAD panel. The SEAD reference panel is now integrated into an imputation server with user-friendly website interface for public use (https://imputationserver.westlake.edu.cn/).

### SEAD panel improves precision in detecting rare variants in South Asian population

To evaluate the imputation performance of SEAD panel in South Asian population, we generated three datasets from UK biobank samples with different proportion (50-70%, 70-90% and >90%) of SAS ancestry composition calculated by Rye software(40) (Figure 1 Step2). We revealed that the SEAD panel consistently exhibited the highest proportion of well-imputed low-frequency sites (Rsq>0.8 & MAF<5%) across all ancestry gradients, particularly in the >90% group (Figure 2A). In contrast, the proportion of well-imputed sites in TOPMed imputation decreased when the SAS ancestry components increased, and fell to less than half from 50-70% group to >90% group (Figure 2A). Similarly, the imputation performance of the ChinaMAP panel declined with increasing South Asian ancestry. The proportion of high quality sites showed little variation across the gradients in 1kGP imputation (Figure 2A and Supplementary table 5). These analyses suggested that despite its large sample size (>130,000), the TOPMed panel performed less effectively in South Asian populations, whereas SEAD panel indicated its superior performance.

To further access the imputation accuracy across Asian populations, we segmented 197 Central and South Asian samples from the HGDP and extracted ASA array sites on chromosome 2 from them to create a pseudo array, and then calculated concordance rate between the imputed and WGS data (Figure 2B and 2C, Supplementary table 6). Compared to the other three panels, the SEAD panel demonstrated higher concordance rate for heterozygote and homozygote genotypes, while the ChinaMAP panel exhibited the lowest concordance (Figure 2B), with 1kGP and TOPMed panels in the middle. Similar results were observed for specificity and precision (Figure 2C). In order to further refine the advantage of each panel, we looked at the performance in each population in Central and South Asian samples in HGDP. The results showed that the SEAD panel consistently demonstrated the highest imputation precision across most populations, often exceeding 0.94 (Supplementary figure 5). For MAF < 0.05 variants, the SEAD panel showed a distinct advantage over the TOPMed and ChinaMAP panels (Figure 2D and Supplementary table 7). The ChinaMAP panel consistently exhibited the lowest accuracy, with median accuracy ranging between 0.7 and 0.8 for low-frequency and rare variants across most populations, reflecting its poorer imputation quality (Figure 2D and Supplementary table 7). All these results suggested that SEAD panel showed optimal performance in South Asian populations.

### Imputation performance in East Asia population

We further evaluated the imputation accuracy of the 1kGP, TOPMed, ChinaMAP and SEAD panels in East Asian populations (Figure 1 Step2). Initially, we extracted WGS data from 223 East Asian samples in the HGDP dataset and created a pseudo array by extracting ASA chip SNPs. The results showed that the peak of SEAD in the density plots were higher than ChinaMAP, indicating that the distribution of concordance rate was more concentrated (Figure 3A and Supplementary table 8), while the ChinaMAP panel had the highest heterozygote and homozygote genotype concordance rates. Concordance rates for the TOPMed and 1kGP panels were lower than those of the two Asian panels (Figure 3A and Supplementary table 8). As for each East Asia population, the SEAD panel outperformed both TOPMed and ChinaMAP panels in Cambodian (which is a South East population) and had similar accuracy to the ChinaMAP panel in the Japanese population (Supplementary Figure 6). In other East Asian populations, the ChinaMAP panel consistently showed higher accuracy than the SEAD panel, while TOPMed consistently had the lowest accuracy (Supplementary Figure 6).

Due to the limited sample size in HGDP East Asian samples (only 223 individuals), it is unable to reflect the imputation performance in MAF (minor allele frequency) bins. We then imputed WBBC-chip data for 5,679 Han Chinese samples using the four panels. The results showed that the ChinaMAP panel consistently demonstrated the highest accuracy (mean Rsq) across all MAF bins, while the SEAD panel outperformed TOPMed and 1kGP panels (Figure 3B and Supplementary Table 9). In terms of the number of well-imputed sites (Rsq>0.8), the count of the sites for SEAD grew closer to that of the ChinaMAP panel, as the MAF increased (Figure 3D). Additionally, we evaluated the imputation quality by comparing the concordance rate in the 179 samples who had both WBBC-seq and WBBC-chip data, where taking the sequencing data as “true”. The results showed that the SEAD and ChinaMAP panels achieved similar and higher homozygote and heterozygote concordance rates, with the lowest concordance rate for the TOPMed panel (Figure 3C and Supplementary Table 10). The similar results were observed for non-reference allele sensitivity and precision (Figure 3D and Supplementary Table 10). These results indicated that the concordance rates of the SEAD and ChinaMAP panels were comparable, whereas the imputation performance of TOPMed panel was inferior to that of the Asian panels.

### Combined panel vs meta-imputation

We compared the performance of meta-imputation of four panels (WBBC-seq, 1kGP, SG10K, GAsP) with the combined SEAD panel (Figure 1 Step2). Across the seven MAF bins, the number of well-imputed loci obtained through SEAD imputation consistently exceeded that of meta-imputation, particularly for rare/low-frequency variants (Figure 3E and Supplementary Table 11). Furthermore, the Rsq values achieved by SEAD imputation were higher in each MAF bin compared to meta-imputation, with an increase of approximately 0.2 observed in the 0.1%-1% MAF region (Figure 3E and Supplementary Table 11). Additionally, we calculated the proportion of well-imputed loci relative to the total number of loci within each MAF bin, where SEAD also demonstrated a clear advantage (Supplementary Figure 7). These results suggested that the combined SEAD panel consistently showed significantly higher well-imputed counts and accuracy across the seven MAF bins compared to the meta-imputation, indicating that the imputation with combined panel is superior to meta-imputation.

### Employment of the SEAD panel in bone mineral density GWAS analysis

After imputing WBBC-chip data (5,369 samples) with SEAD panel, we conducted GWAS analysis on two BMD traits (total hip and femoral neck BMD) (Figure 1 Step3). These two BMD traits were highly correlated in either Pearson correlation (*r* = 0.918) or genetic correlation (*r* = 0.932, SD = 0.025). With genome-wide significance threshold (*P* < 5×10^−8^), two common loci (MAF > 0.01) that have been previously reported in GWAS were identified to be associated with both BMD traits, with the top signals at chr1:rs9659023 (an intronic variant of *FMN2*, this locus was reported to be associated with heel (60) and total body BMD (61)) and at chr6:21392233:GA:G (near *SOX4*, this locus was reported to be associated with heel (60) and FN BMD (13)) (Supplementary Figure 8).

For the relatively rare variants (0.001 < MAF < 0.01), we utilized two analytical strategies: single-variant test with PLINK and variant-set analysis via the STAARpipeline (incorporating both sliding-window and gene-centric methods). In the single-variant test analysis, we prioritized the variants with small P-value and with replication (see Methods), and identified 106 suggestive variants for Hip BMD (Supplementary Table 12). Among these variants, 68 (64.1%) were annotated to *SNTG1* gene by ANNOVAR (Supplementary Table 12). Of which, four rare variants (rs60103302, rs61260287, rs60600379 and rs57319781), in complete linkage disequilibrium (LD r^2^=1.00), were identified for Hip BMD at near genome-wide significance level (*P* = 1.67×10^−7^, MAF = 0.0092), these variants located in the intergenic region near *SNTG1* on chromosome 8 (Figure 4A and 4F and Supplementary Table 12). As for FN BMD, 1 out of the 35 suggestive variants were annotated to *SNTG1* gene (Figure 4C and 4F, and Supplementary Table 12). In the variant-set analysis, we first implemented a sliding window of 2kb in STAAR-O test and adhered to the previous “discovery and replication” strategy. The results showed that 5 clustering regions were identified as the suggestive signals for Hip BMD (Figure 4B, Supplementary figure 9 and Supplementary Table 13). In the cluster on chromosome 8 (chr8:49873459-49958458), a total of 30 windows were suggested, of which, 3 windows annotated near *SNTG1* were discovered at bonferroni-corrected level *P* = 3.59×10^−8^ (0.05 divided by 1,392,296 windows), with top window at chr8:49903244-49905243 (*P* = 9.08×10^−9^) (Figure 4B, Supplementary figure 9 and Supplementary Table 13). Moreover, in the annotation-based gene-centric analysis, of the 7 non-coding categories of *SNTG1* gene, 8 categories showed suggestive signal for Hip BMD, with the top signal at promoter region (*P_promoter_CAGE_* = 5.72×10^−8^) (Figure 4E, Supplementary figure 9 and Supplementary Table 14), showcasing at least one annotation reaching the genome-wide bonferroni-corrected level *P* = 3.57×10^−7^ (0.05 divided by 20,000 genes then divided by 7 categories). In summary, *SNTG1* locus was highlighted in both analytical strategies with the strictest threshold applied to Hip BMD, and also achieved a suggestive level in FN BMD (*P_single_variant_* = 7.79×10^−6^, *P_slide_window_* = 1.34×10^−6^, *P_gene_centric_* = 5.71×10^−6^) (Figure 4C, 4D, 4E, Supplementary figure 9, Supplementary Table 13 and 14),

We compared the frequency distribution of the suggestive rare signals identified in *SNTG1* locus (72 SNPs) with the gnomAD and TOPMed databases (as they both have very big sample size) across various populations (Figure 4H, Supplementary Table 15). The *SNTG1* variants were predominantly rare in the Non-Finnish European (NFE: 34,029 samples) and South Asian (SAS: 2,419 samples), while they were more common in the African/African American (AFR: 20,744 samples) and TOPmed (dominantly European population), highlighting the population-specific patterns of this region. The MAF of *SNTG1* variants imputed by SEAD panel most closely aligned with the MAF value in the East Asian (EAS) population (2,604 samples) within gnomAD v3 (Figure 4H), validating the appropriateness of SEAD panel to identify Asian-specific associated variants.

### SEAD panel vs other panels in detecting Asian-specific *SNTG1* locus

To test if other panels are also capable of identifying *SNTG1* locus, we imputed WBBC-chip data using the TOPMed, 1kGP, SG10K, GAsP, ChinaMAP and WBBC-seq panels and performed association analyses on hip BMD. We assessed the imputation quality (Rsq) of associated variants (meta P-value<1e-4) for the WBBC-chip dataset imputed with each panel within the *SNTG1* region chr8:49,400,000-50,400,000 (Figure 4I). The results showed that the mean imputation Rsq for the SEAD, ChinaMAP, WBBC-seq and SG10K panels exceeded 0.9, while for the TOPMed and 1kGP panels, it ranged between 0.6 and 0.7 (Figure 4I and Supplementary Table 16). After applying a less strict threshold of Rsq>0.5, the association signals at the *SNTG1* locus were detectable in the WBBC-chip dataset imputed with all Asian panels (Supplementary figure 10), but not with the TOPMed and 1kGP panels (Supplementary figure 11). As for the most significant SNPs, the SEAD, ChinaMAP, WBBC, and SG10K panels demonstrated the same top SNP (rs60103302), with comparable effect size, MAF and genotype counts, except for the GAsP panel (Figure 4G). For the TOPMed and 1kGP panels, the most significant SNPs showed p-values greater than 1×10^−4^ with poor Rsq (Figure 4G and Supplementary figure 11).

These results demonstrated that, although TOPMed possesses largest-scale reference haplotypes and contains a subset of Asian samples, it is not efficient to impute data in Asian population, emphasizing the significance of population-specific reference panel.

### Preliminary in-vitro analysis of *SNTG1* gene on osteogenesis

To determine whether the associated SNP would affect the transcriptional activity of *SNTG1* gene, we selected the intronic SNP rs111829635 (instead of the intergenic top SNP rs60103302) to perform the dual-luciferase assays in two cell lines, 293T and MC3T3-E1. 483-bp fragments containing the rs111829635 were inserted into a luciferase reporter plasmid with respect to a minimal promoter. It was observed that the intronic SNP rs111829635 C-T mutation significantly increased luciferase activity in both 293T cells and MC3T3-E1 cells (Figure 5A), suggesting that the identified locus would alter the *SNTG1* gene expression.

Moreover, we utilized the CCK-8 proliferation assay to examine the effects of SNTG1 overexpression in mouse preosteoblast MC3T3-E1. The results showed that the absorbance at OD 450 nm was decreased after *SNTG1* overexpression, and cell density was also decreased (*P* <0.05, Figure 5B and Figure 5C), revealing that overexpression of *SNTG1* led to a significant reduction in cell proliferation. At the 72 hours, a noticeable decrease in the expression of osteogenic marker genes (*RUNX2*, *COL1A1* and OCN) confirmed that *SNTG1* inhibited the differentiation of preosteoblast MC3T3-E1 cells (*P* <0.05, Figure 5D). Furthermore, we measured alkaline phosphatase (ALP) level, a marker of osteoblast activity, which showed a significant reduction after six days of *SNTG1* overexpression (Figure 5D). Overall, these results supported the notion that *SNTG1* overexpression might inhibit human osteogenic proliferation and differentiation, highlighting its potential role in osteogenesis.

## Discussions

In this study, we integrated whole-genome sequencing data from SG10K, GenomeAsia, WBBC, and 1kGP-Asian to create a combined reference panel for genotype imputation, the South and East Asian reference Database (SEAD). It comprised a diverse range of populations across Asia, including 11 populations from GenomeAsia, one population from WBBC, three populations from SG10K, and 8 populations from 1kGP-Asian. With a sample size of 22,134 haplotypes, the SEAD panel stands as one of the most comprehensive panels in terms of coverage across Asia. We compared the concordance rate of the genotypes imputed from the SEAD panel across Asian populations with the TOPMed, ChinaMAP and 1kGP panels, and suggested that the SEAD panel performed the best for the rare variants imputation in South Asian populations. Meanwhile, the concordance rates of the SEAD and ChinaMAP panels were comparable for imputing East Asian populations, whereas the imputation performance of TOPMed panel was inferior to that of the Asian panels. Finally, we applied the SEAD panel to the bone mineral density GWAS analyses in WBBC-chip data, and identified an Asian specific rare locus, *SNTG1*, that was not reported even in the large biobank-scale GWAS.

Asia, being the largest and most populous continent worldwide, boasts a wealth of human genetic resources. However, most of the whole genome-sequencing (WGS) efforts were carried out in Caucasian populations in the last decade (17). In our previous study, we conducted a thorough evaluation and discussion on the imputation of rare variants, highlighting the necessity of constructing a haplotype reference panel for Asian populations (10, 62-64). In recent years, there has been a notable surge WGS data across Asia, particularly in regions such as Japan (18), Singapore (20), Korea (65) and China (22, 24, 66). Despite the abundance of WGS projects, large-scale reference panel with broad geographical coverage across Asia is still needed. The accumulation of such datasets from diverse Asian populations presents a unique opportunity for amalgamating multiple WGS datasets into a singular, more comprehensive, and expansive reference panel, which would encapsulate a broader spectrum of genetic diversity, thereby enhancing its utility for human genetic study in Asian populations. The recently published Northeast Asian Reference Database, with over half of the samples deriving from Japan and Korea, represented populations from Northeast Asia (67). In our study, we conducted joint-calling between WBBC-seq and 1kGP-Asian, followed by mutual imputation with SG10K and GAsP to construct the South and East Asian reference Database (SEAD) panel, covering the widest geographical area in Asia, with the majority of its population originating from South and East Asia.

The state-of-the-art imputation reference panel released by the TOPMed includes a diverse range of populations, such as African Americans, Hispanic/Latino, Asian and Caucasian populations. Despite the large and diverse samples in the TOPMed panel, imputation for some populations, notably those from Asia, Oceania and the Pacific, would not fully benefit from it (30). In our study, we observed that the TOPMed panel performed less effectively than the SEAD panel in both South Asian and East Asian populations, but better than the 1kGP panel in East Asian populations, particularly for the low-frequency variants. The ChinaMAP panel contains the largest number of Chinese samples (23), but it performed the worst in all the Central and South Asian populations, in which the SEAD panel performed the best. However, benefiting from the large sample size, the ChinaMAP panel indicated its superior performance in Chinese population. Prospectively, we will update the SEAD panel in the future with additional East Asian samples to improve its imputation efficiency in the corresponding population. All in all, the SEAD reference panel demonstrated significant advantages in genotype imputation for South Asian populations and exhibited good performance in imputation accuracy for East Asian populations. On the other hand, meta-imputation could integrate the imputation results generated using separate reference panels into a consensus dataset (54). And Yu et al demonstrated that this method might generate comparable accuracy to the imputation with a combined reference panel (54). However, based on our assessment, the combined SEAD panel consistently showed significantly higher well-imputed counts and accuracy across each MAF bin compared to meta-imputation with separate reference panels. In addition, meta-imputation might not improve imputation accuracy for underrepresented populations (30).

Besides the systematical and comprehensive imputation evaluation for the panels, we successfully applied the SEAD panel to a GWAS analysis for BMD traits at hip and femoral neck (FN). These two traits were highly correlated in either phenotypic or genetic correlation. Therefore, the association signals reported in both traits would reduce the random error of the results. Through both single-variant association test and variant-set analysis, we detected that rare variants near *SNTG1* gene were associated with hip BMD. The *SNTG1* gene also reached the suggestive threshold in FN BMD analysis using both methods. The gene and variants were not reported even in large-scale biobank GWAS for BMD (60), and the *SNTG1* variants were predominantly rare in the Non-Finnish European and more common in the African/African American and Latino/Admixed American. The *SNTG1* signal for hip BMD was identified in GWAS imputed with all Asian panels, but not the TOPMed and 1kGP panels. Specifically, the rare variant rs60103302 (MAF=0.0092) was identified to be the top signal in imputation with the SEAD panel, as well as the SG10K, WBBC-seq and ChinaMAP panels. However, this SNP had poor Rsq in TOPMed imputation (Rsq=0.4172), and no SNPs showed SNPs showed p-values less than 1×10^−4^. This is because the low-frequency and rare variant associations are more likely to be population specific, which would suffer a relative loss in statistical power in GWASs (30). Therefore, the incapability of TOPMed and 1kGP to impute Asian-specific rare variants further underscores the importance of building Asian-specific panels for the discovery of rare variants unique to Asian populations.

The *SNTG1* (Syntrophin Gamma 1) is identified as a neuronal cell-specific protein, prominently expressed in the Purkinje neurons of the cerebellum and the pyramidal neurons of the hippocampus and cortex(68), suggesting its potential role in neuronal stability and signal transmission within the nervous system (68). The nervous system, particularly through the sympathetic nervous system (SNS), plays a crucial role in regulating bone metabolism. The SNS provides innervation to bone tissue, with the sympathetic fibers directly interacting with osteoblasts and osteoclasts (69). Clinical evidence supported the involvement of the SNS in bone metabolism, linking conditions associated with increased sympathetic activity (such as reflex sympathetic dystrophy) to decreased bone mineral density (70, 71). The *SNTG1* was identified as a candidate gene for idiopathic scoliosis (72–74). A southern Chinese cohort of patients with congenital scoliosis also identified copy number variants of *SNTG1* (75). The underlying pathological mechanisms of idiopathic scoliosis remain unclear, a prevalent theory posits that the primary abnormality is a neurological defect. This defect causes abnormal processing in the central nervous system (CNS), leading to developmental anomalies in the spine (72). Animal models of scoliosis indicated damage to the cerebellum in bipedal rats induced scoliosis (76). Additionally, the SNTG1 protein was involved in the Dystrophin-associated glycoprotein complex (DGC) pathway. The DGC was known to play a significant role in maintaining the structural integrity of muscle fibers (77). Reduction in muscle strength diminished mechanical stimuli to bones, adversely affecting bone remodeling and maintenance. Sarcopenia, characterized by decreased muscle mass and strength, can increase the risk of osteoporosis, particularly in the elderly (78, 79). Further, muscle atrophy could influence bone density through hormonal and inflammatory pathways (80, 81). In our study, we suggested that overexpression of *SNTG1* inhibited the proliferation and differentiation of preosteoblasts. Given that osteoblasts are pivotal in orchestrating bone formation and osteogenesis, a reduction in their proliferation and differentiation capacity translates into a diminished osteoblast pool. This, in turn, leads to a decrease in bone formation and the overall process of osteogenesis. This observation aligned with the direction of effect on BMD in our GWAS results.

In summary, we constructed a haplotype reference panel for Asian population (SEAD panel) with the largest number of samples and the most abundant population diversity in Asia. The reference panel demonstrated excellent performance on imputing East Asian and Central and South Asian populations, especially in detecting rare variants. We provided this optimal imputation service online for free (https://imputationserver.westlake.edu.cn/) for genetic studies in Asian populations. By applying the SEAD panel to impute the genotyping array data in Chinese population, we, for the first time, successfully identified rare variants near *SNTG1* gene showing association with bone mineral density.

## Data Availability

GWAS summary statistics for hip and femoral neck (FN) bone mineral density (BMD)from Westlake biobank for Chinese

https://wbbc.westlake.edu.cn/downloads.html

## Data availability

The SG10K (dataset ID:EGAD00001005337 on https://ega-archive.org/) and GAsP (http://portal.us.medgenome.com/GA100K_phase1_12102019/) datasets were obtained by applying to the consortium. The high-coverage 1kGP dataset with an average depth of 30×, based on the GRCh38 assembly, were downloaded from the https://www.internationalgenome.org/data-portal/data-collection/30x-grch38. The CRAM files for Asian ancestry samples (EAS and SAS) from 1kGP were downloaded from here (ftp://ftp.sra.ebi.ac.uk/vol1/run/ERR323/). The SEAD reference panel is now integrated into an imputation server with user-friendly website interface for public use (https://imputationserver.westlake.edu.cn/). Users can register and create imputation jobs freely by uploading their bgzipped array data (VCF-formatted) to the server under a strict policy of data security. The UK Biobank data was directly downloaded upon request (Application 41376) (https://www.ukbiobank.ac.uk/). The GWAS summary statistics for the Hip and FN BMD of the WBBC-chip data are fully available at https://wbbc.westlake.edu.cn/downloads.html.

## Acknowledgements

We thank the “SG10K_Pilot Investigators” for providing the SG10K_Pilot data (EGAD00001005337). The data from the “SG10K_Pilot Study” reported here were obtained from EGA. This manuscript was not prepared in collaboration with the “SG10K_Pilot Study” and does not necessarily reflect the opinions or views of the “SG10K_Pilot Study”. We also thank the “Genome Asia 100K consortium” for providing the “GenomeAsia pilot project” (EGAS00001002921). We thankfully acknowledge the High-performance Computing Center at Westlake University.

## Funds

This work was supported by the National Natural Science Foundation of China (#82370887), the Chinese National Key Technology R&D Program, Ministry of Science and Technology (#2021YFC2501702). And the “Pioneer” and “Leading Goose” R&D Program of Zhejiang (#2023C03164).

## Author contributions

H.-F.Z. conceptualized and designed the study. M.-Y.Y., J.-D.Z., G.T., and W.-Y.B. conducted the data analysis. X.L. conducted in-vitro experiments. S.-H.Y., W.-W.Z., J.-Q.L. and Y. S. conducted the whole sequencing experiments. C.-D.Y., M.-C.Q., Y.-H.F., C.-F.Y., P.-K.C., K.S., S.-R.G., P.-P.Z., P.-L.G., Y.Q., J.-G.T., X.-J.Y., J.-X.G., X.C., M.-M.M., L.-X.L., G.T., S.-Y.X., L.X., F.H., J.-C.L., J.-F.G., B.-S.T., L.Y. and D.K. contributed to the sample collection, processing and preliminary data analysis. J.-J.Y., Y.-H.L. and N.L. designed the online website resource. M.-Y.Y. and J.-D.Z. drafted the manuscript, H.-F.Z. reviewed and edited manuscript. All authors contributed, discussed and approved manuscript.

---

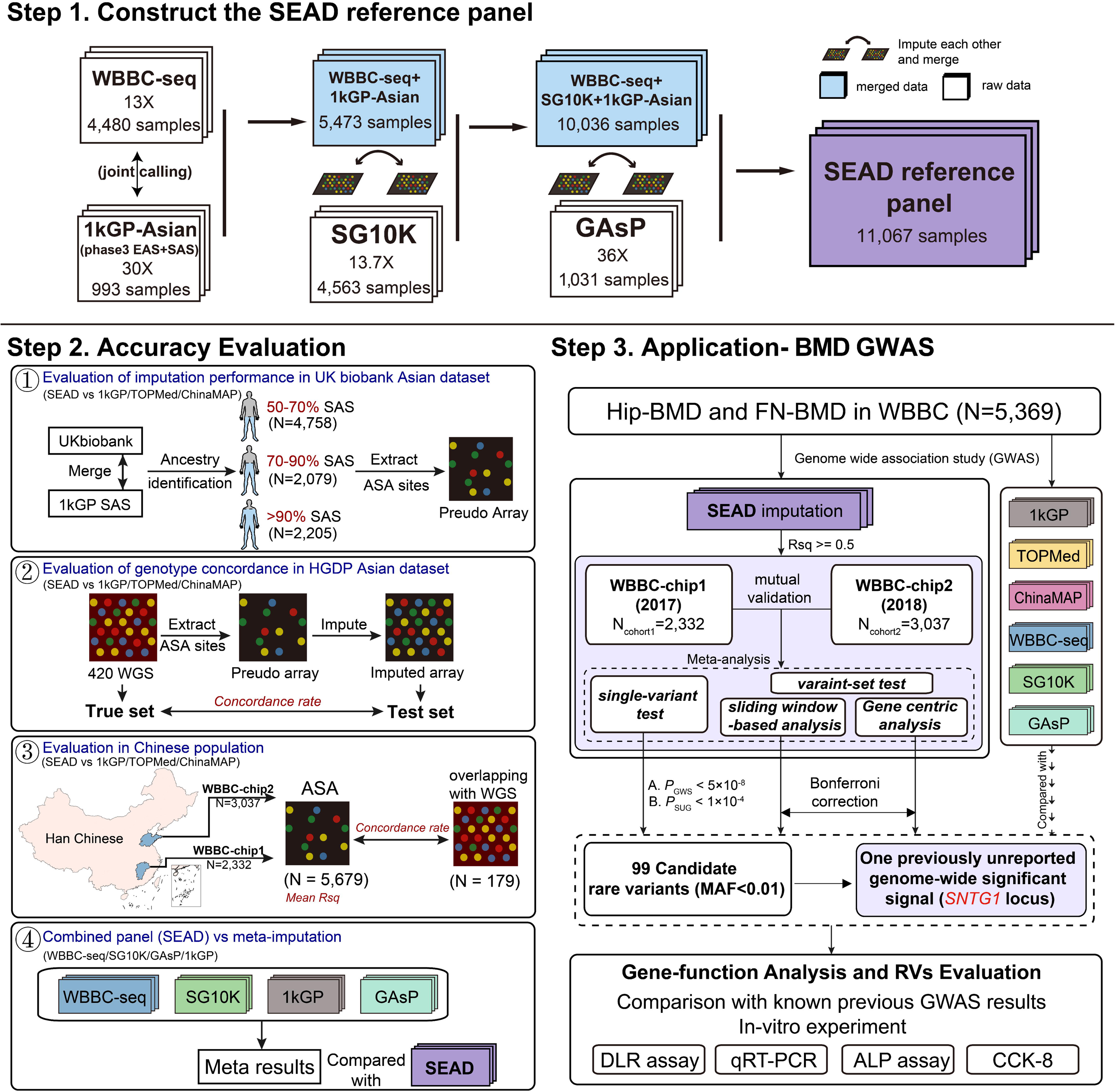

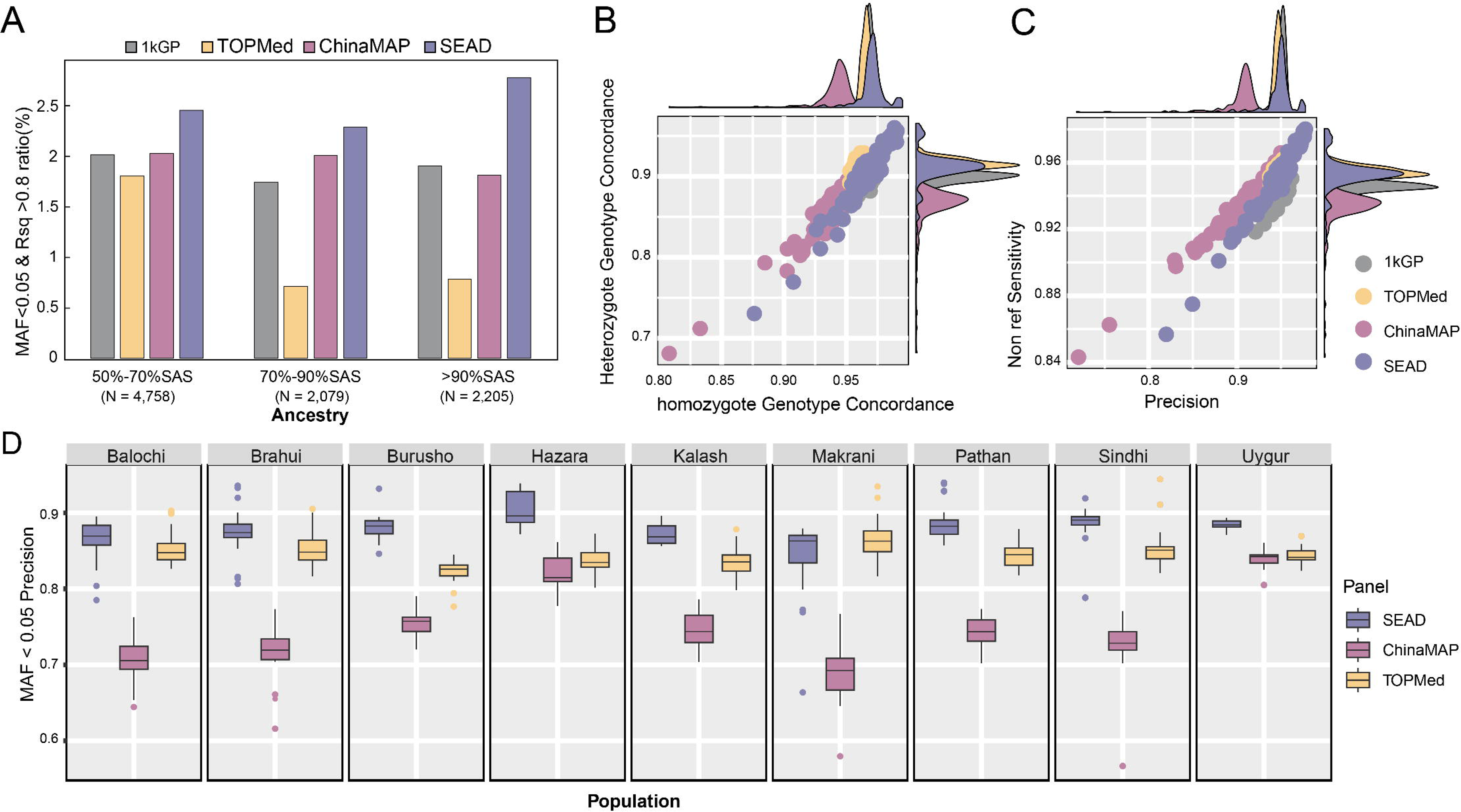

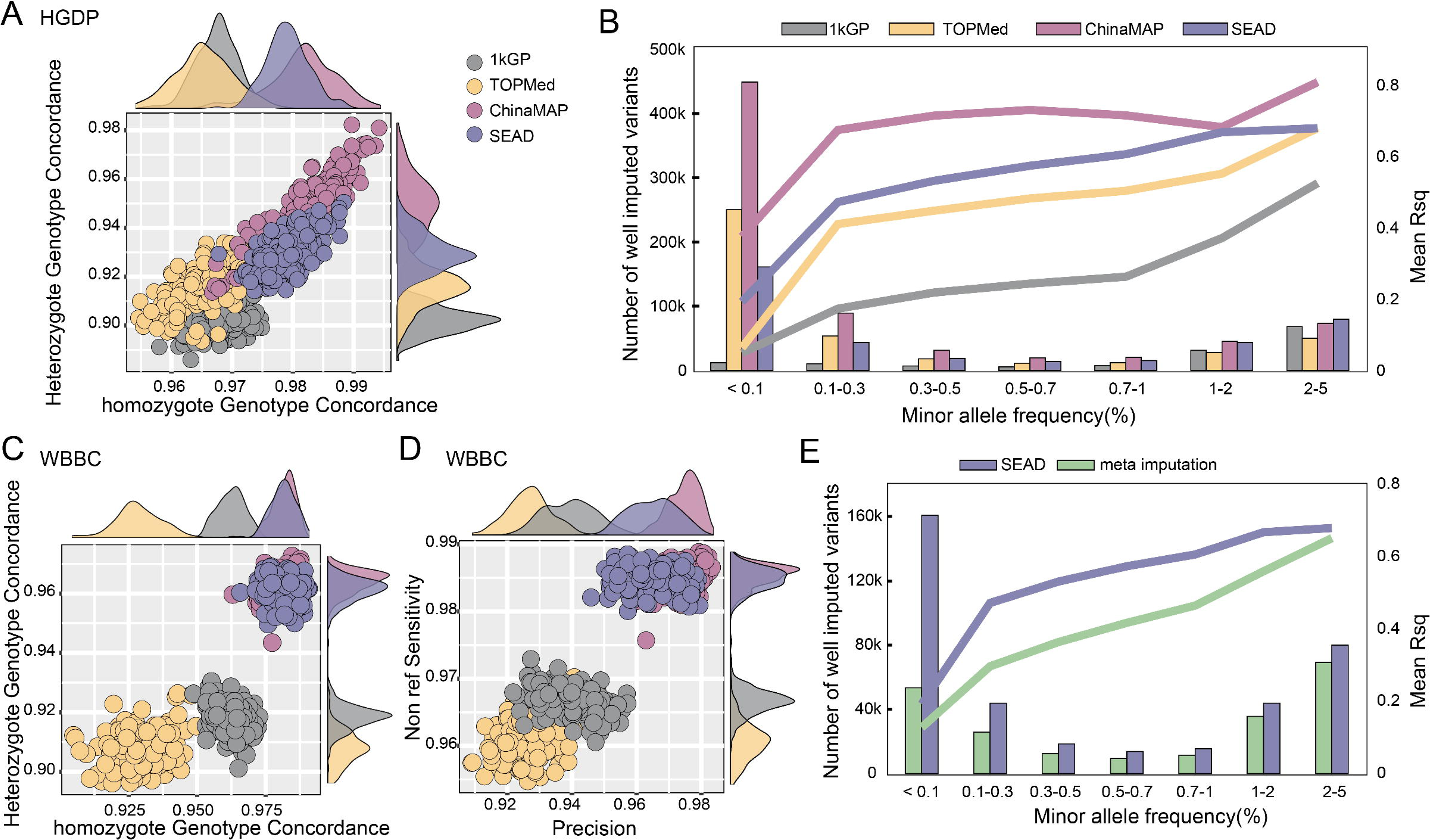

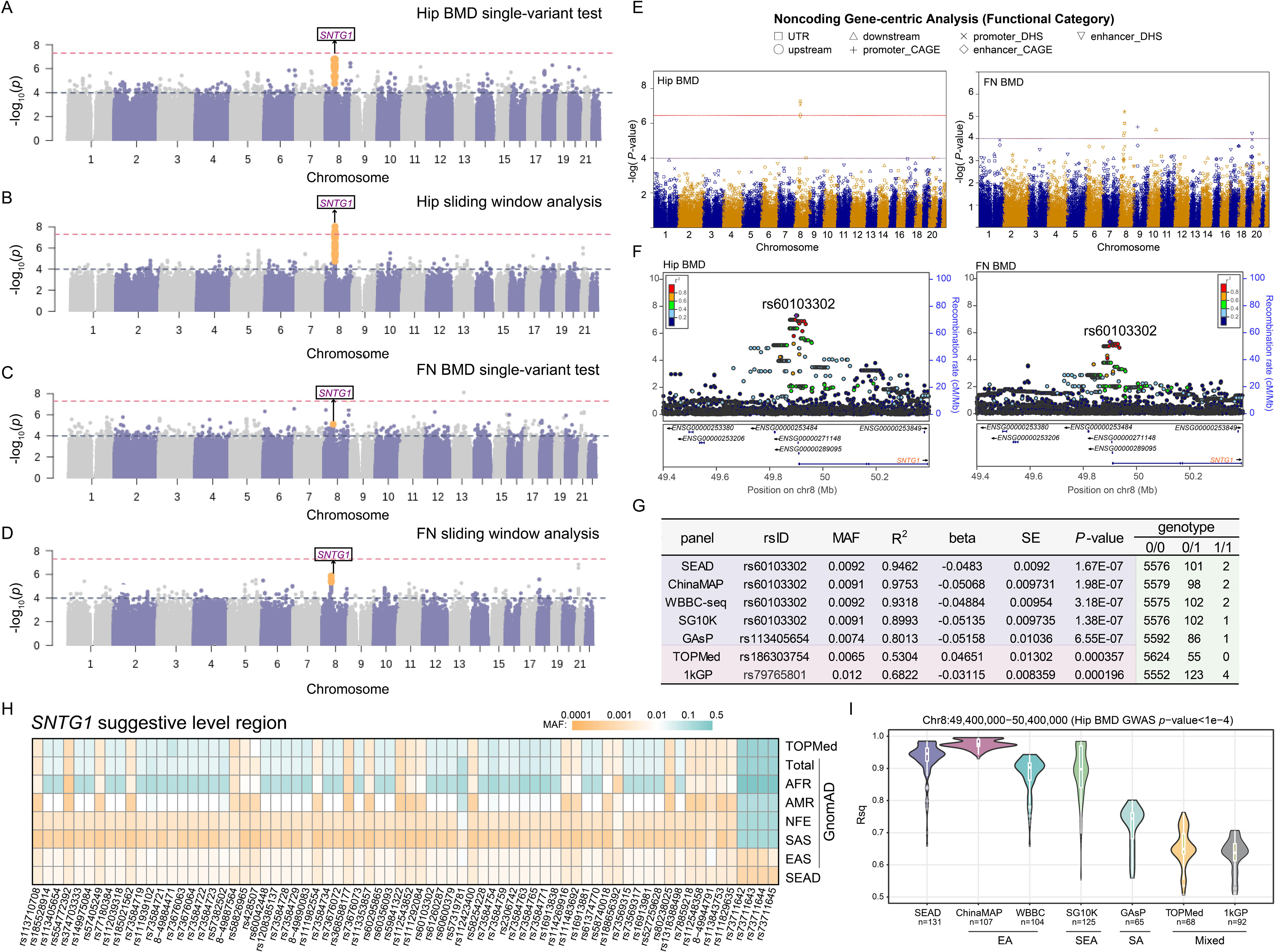

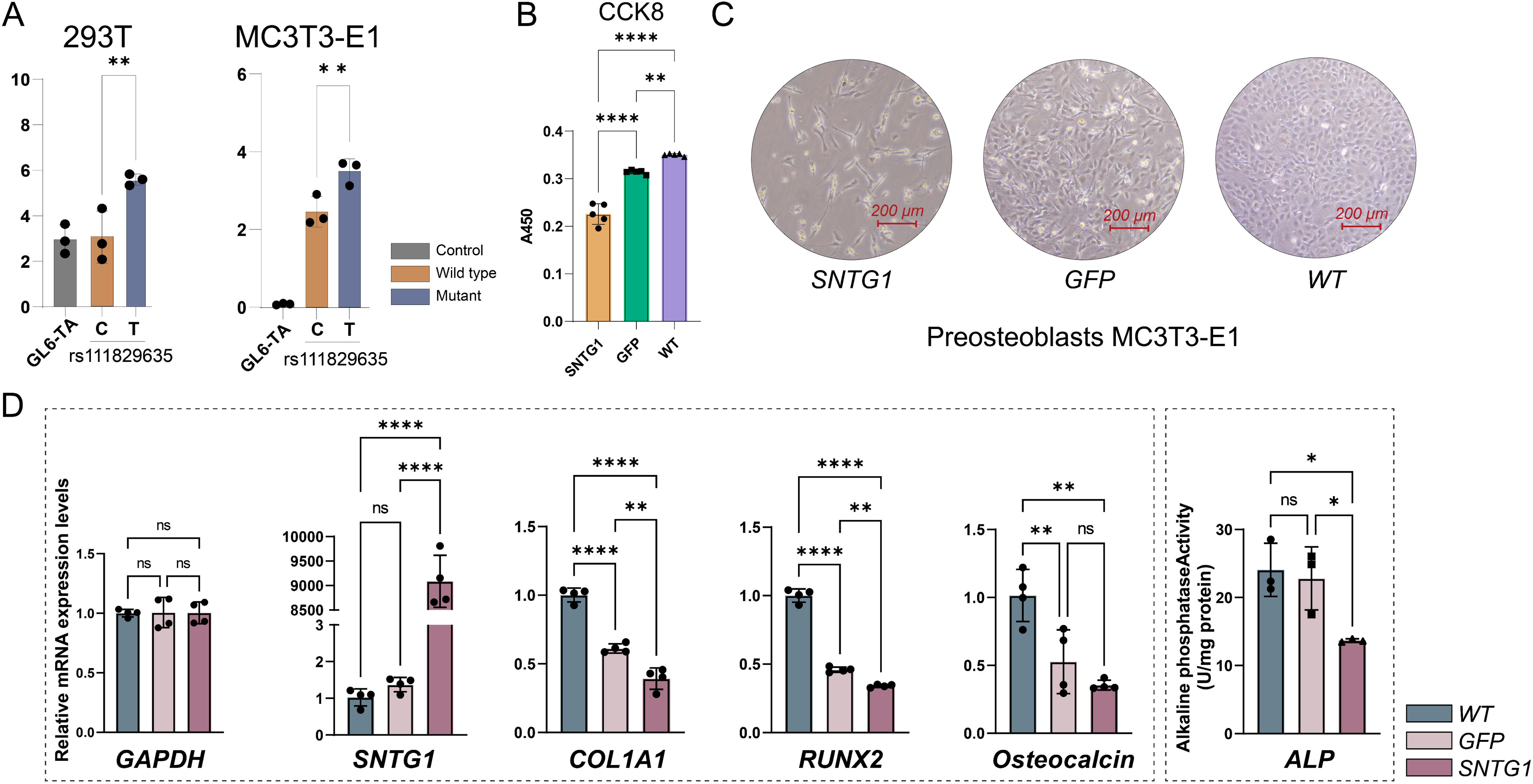

