## Supplementary figure for "SEAD: an augmented reference panel with 22,134 haplotypes boosts the rare variants imputation and GWAS analysis in Asian population": Supplementary_figure_final.docx

**Supplementary figure 1.** Definition of Genotype Concordance, Precision and Sensitivity

0 represents reference allele while 1 represents Non-Reference (NR) allele, dot means allele missing called. For each metrics, the value equals to the corresponding red rectangles divided by all rectangles with blue border.

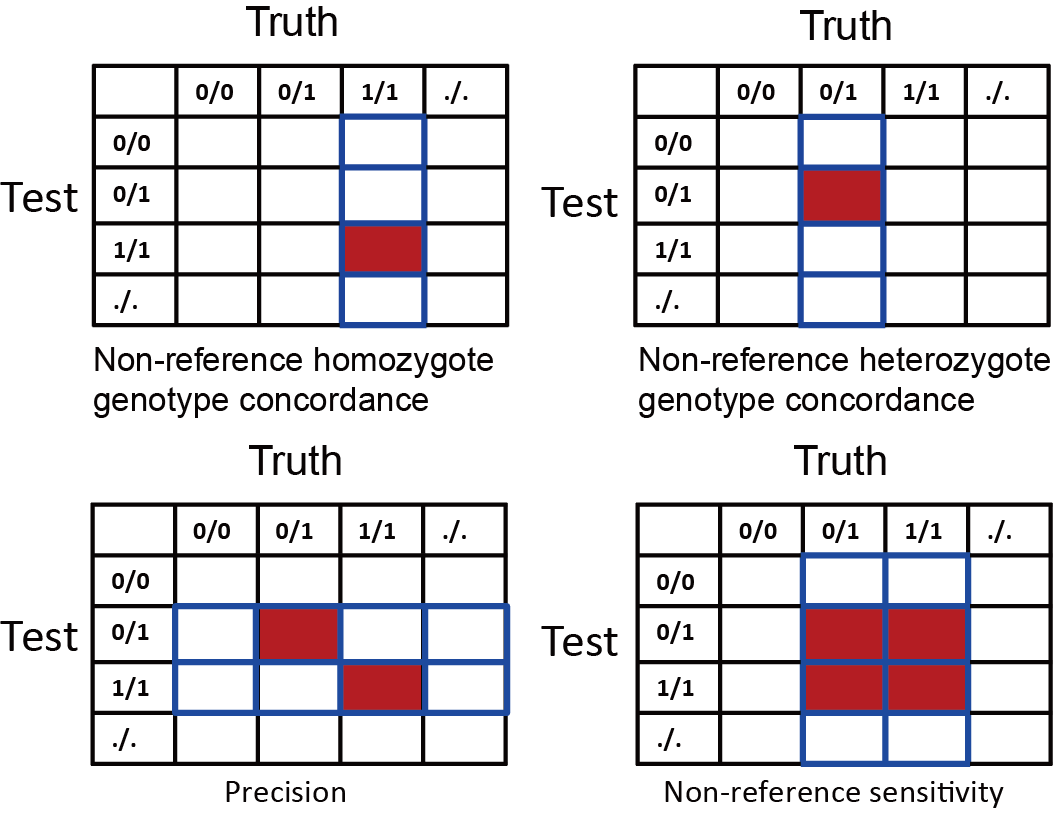

**Supplementary figure 2.** The SNV number and ts/tv ratio of WBBC and WBKG joint calling data.

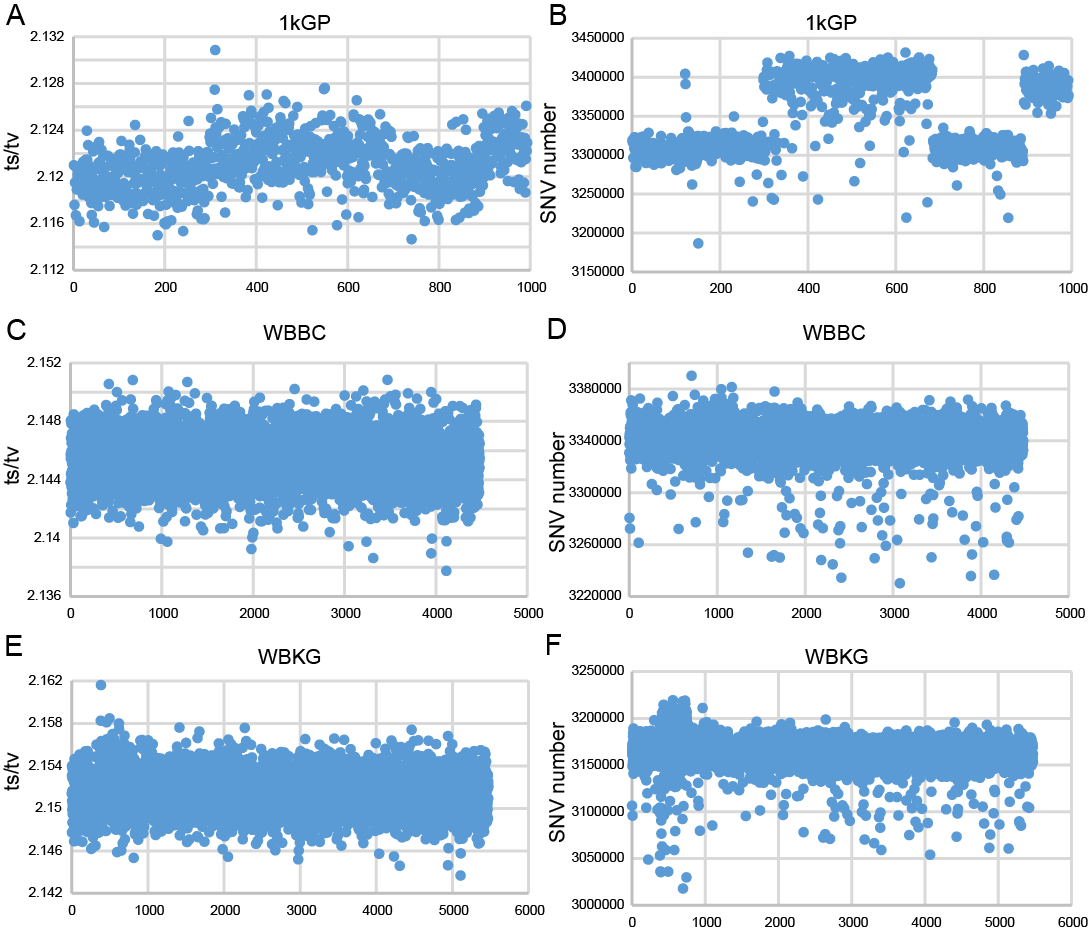

**Supplementary figure 3.** The non ref genotype concordance, heterozygote genotype concordance and homozygote genotype concordance rate of 179 replicate samples (array VS WGS).

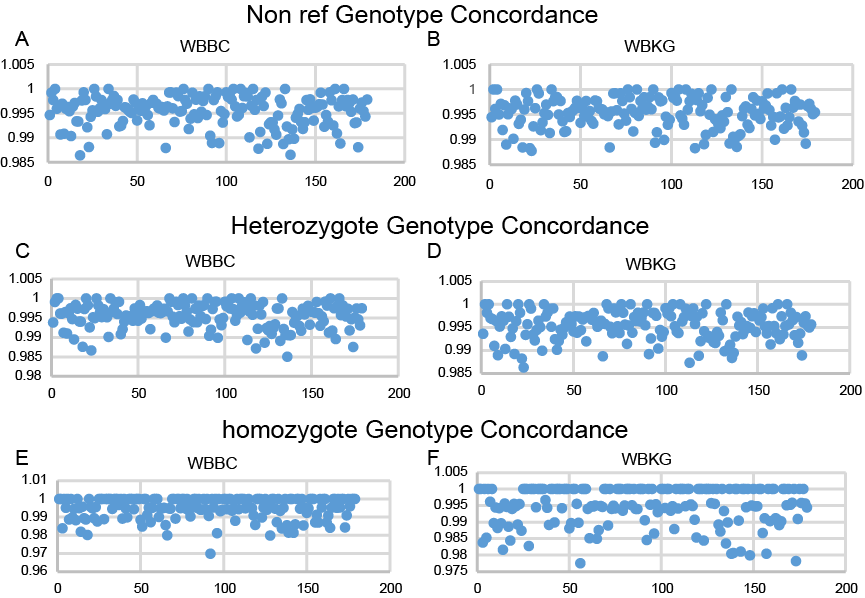

**Supplementary figure 4.** The PCA of SEAD panels with raw variants and GQ > 40, DP > 20 variants.

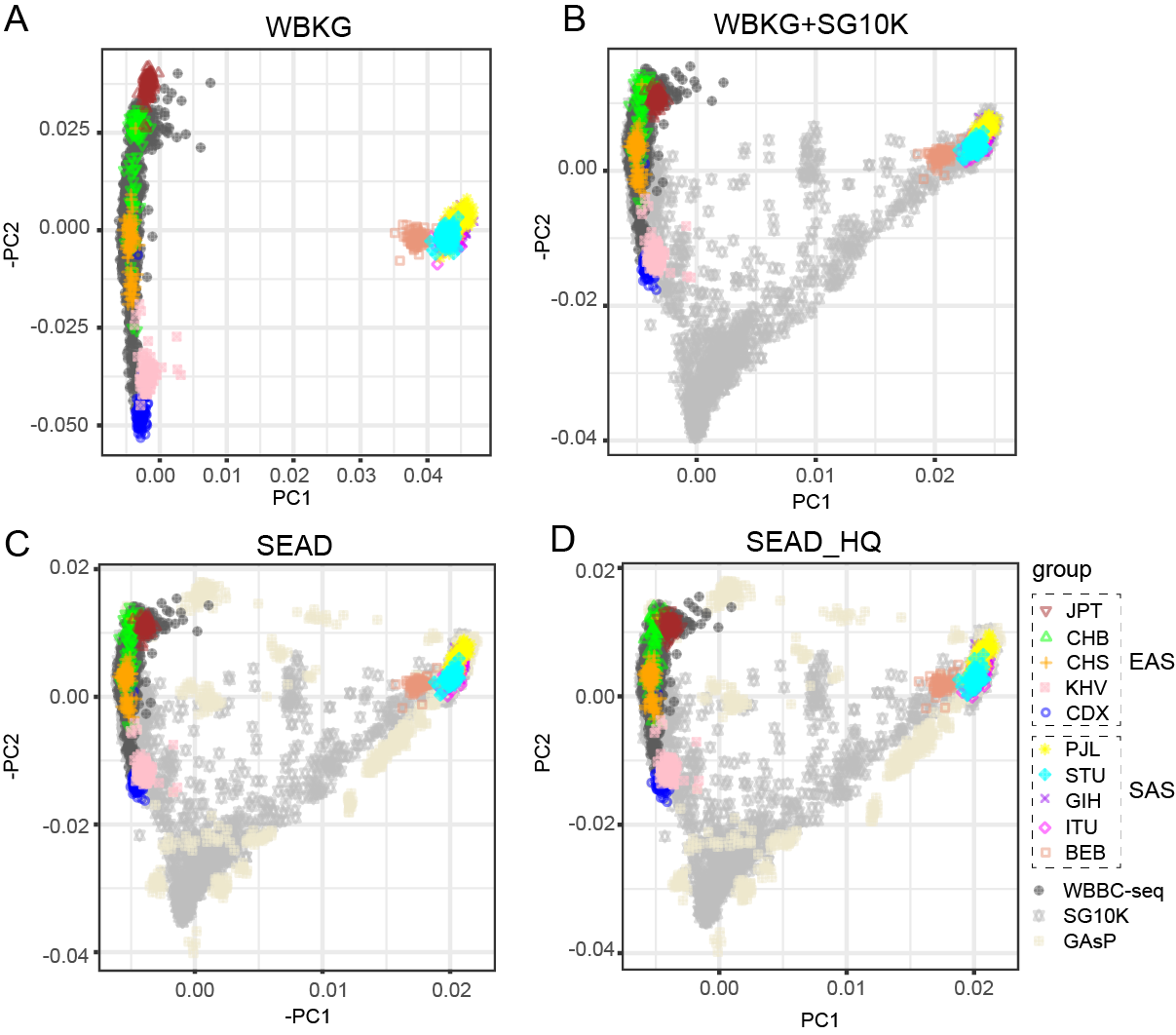

**Supplementary figure 5.** The precision in all variants across 9 Central and South Asian populations in HGDP.

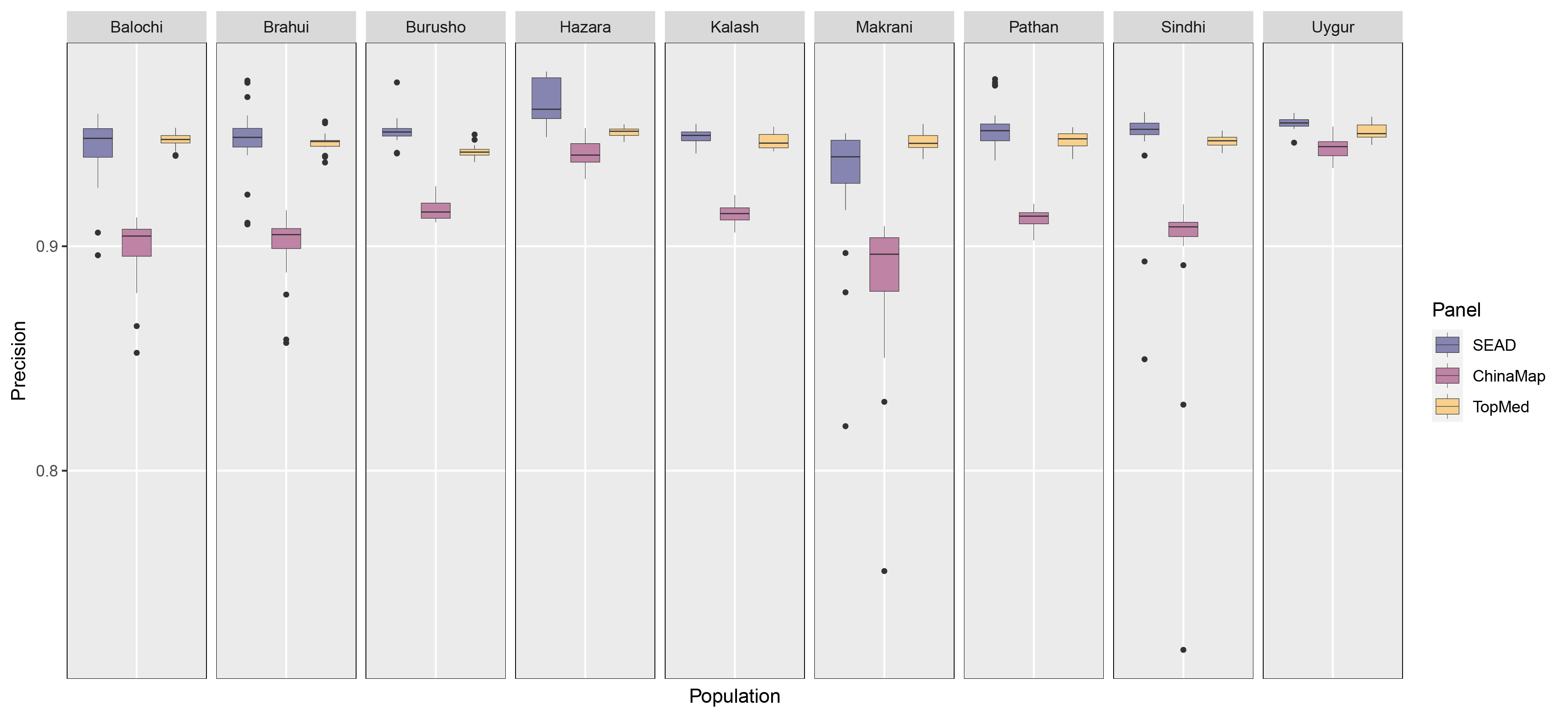

**Supplementary figure 6.** The precision in all variants across 18 East Asian populations in HGDP.

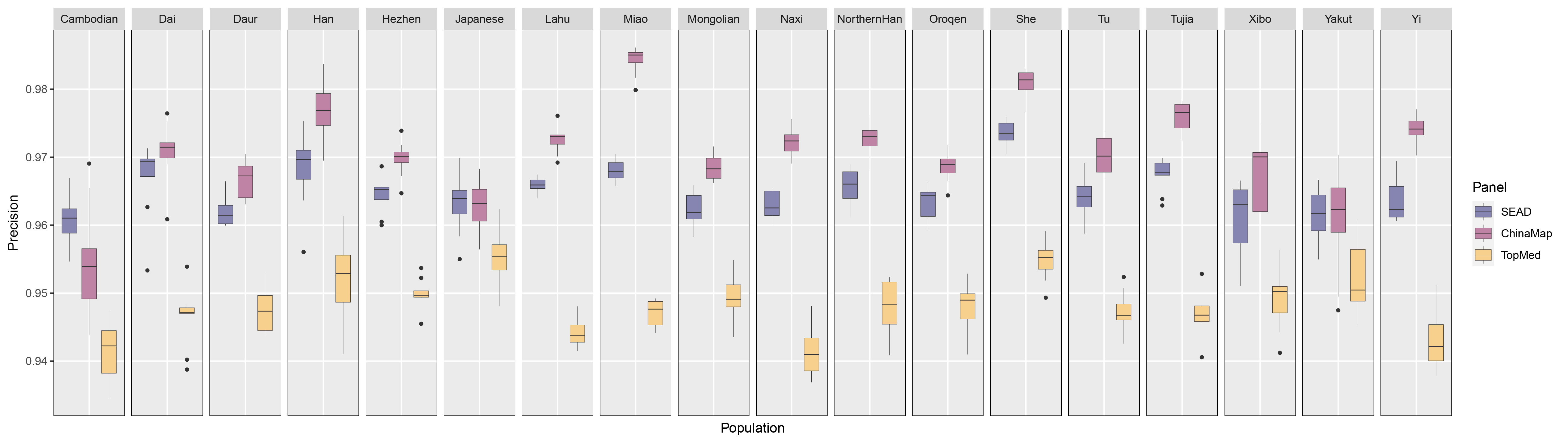

**Supplementary figure 7.** The well imputed number and ratio of meta imputation and SEAD imputation

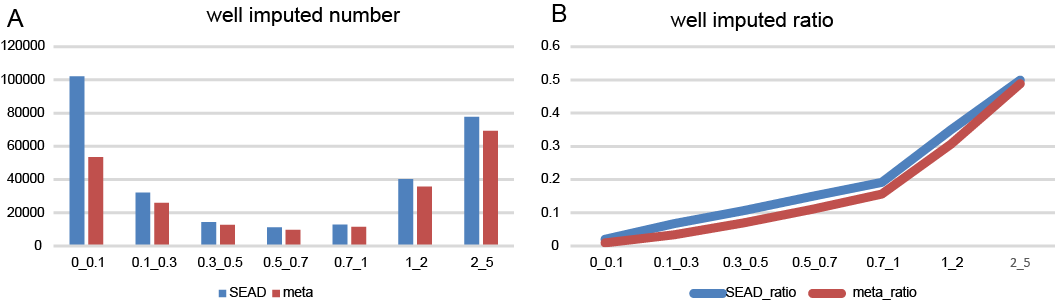

**Supplementary figure 8. Hip and FN BMD GWAS analysis (all variants)**

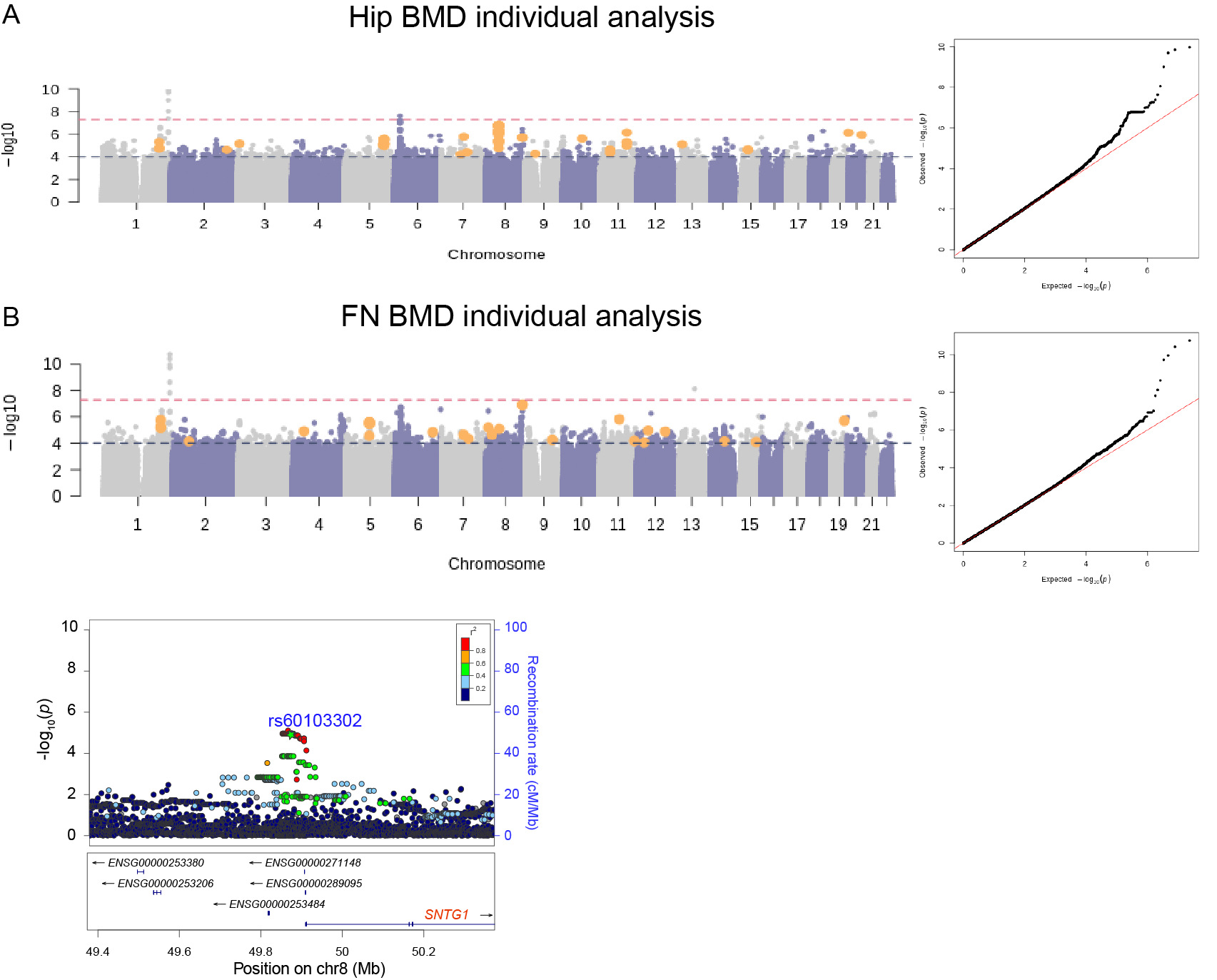

**Supplementary figure 9. QQ plot of low-frequency and rare variants (MAF <= 0.05)**

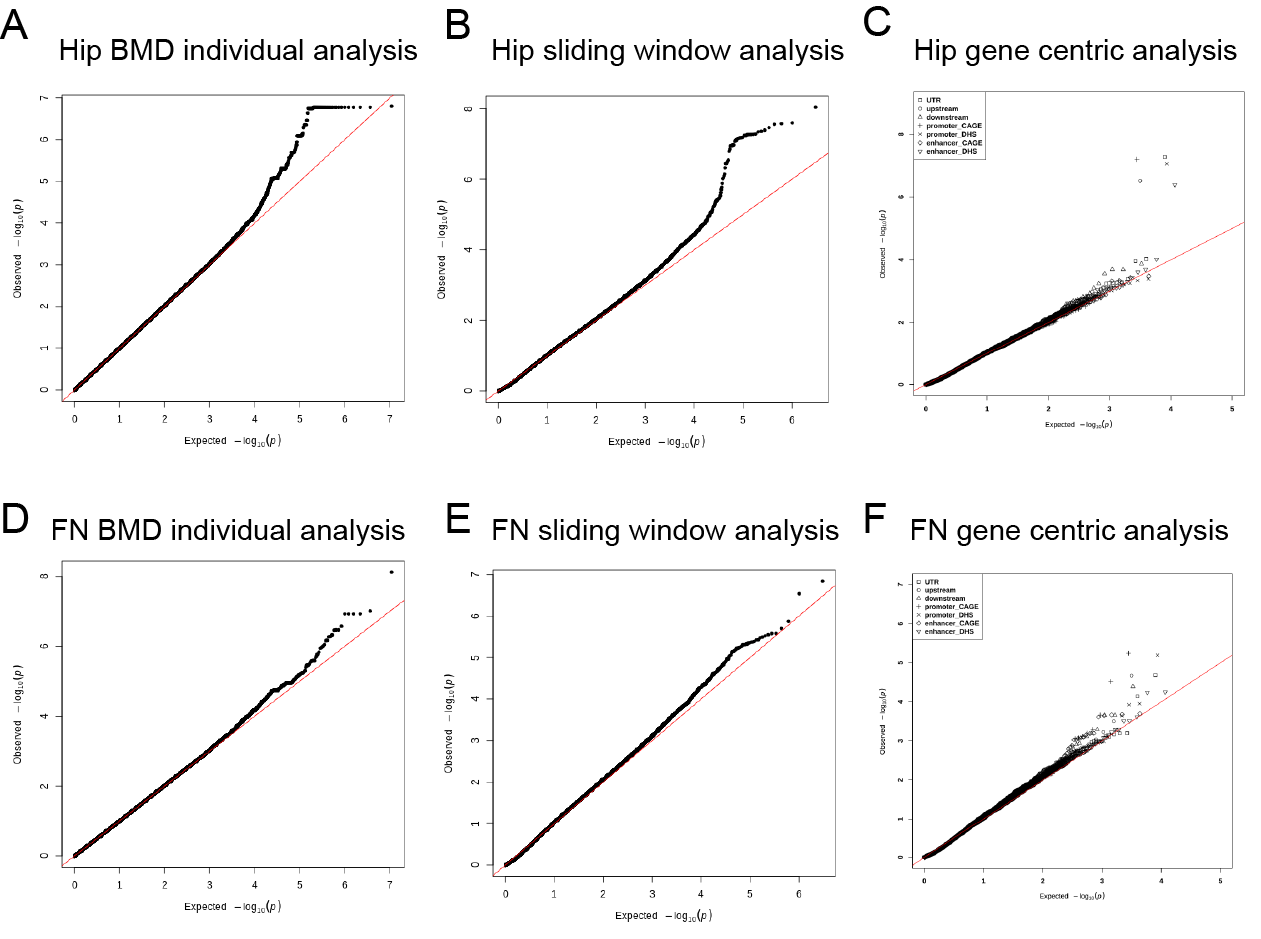

**Supplementary figure 10.** Manhattan and QQ plot with all variants (MAF > 0.001) imputed by ChinaMAP, SG10K, and WBBC panel of Hip BMD using GWAS individual analysis.

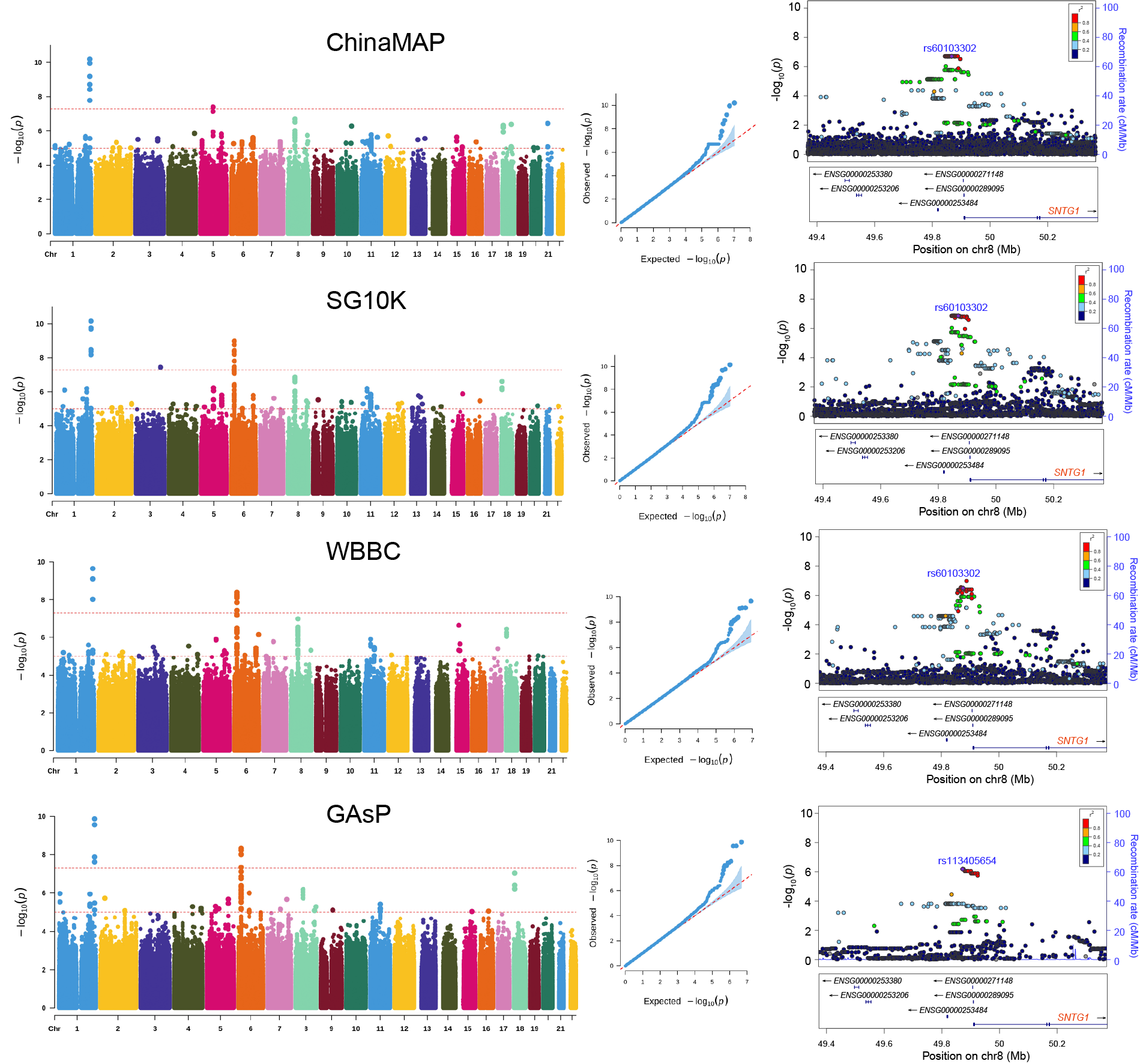

**Supplementary figure 11** Manhattan and QQ plot with all variants (MAF > 0.001) imputed by TOPMed, 1kGP, and GAsP panel of Hip BMD using GWAS individual analysis.

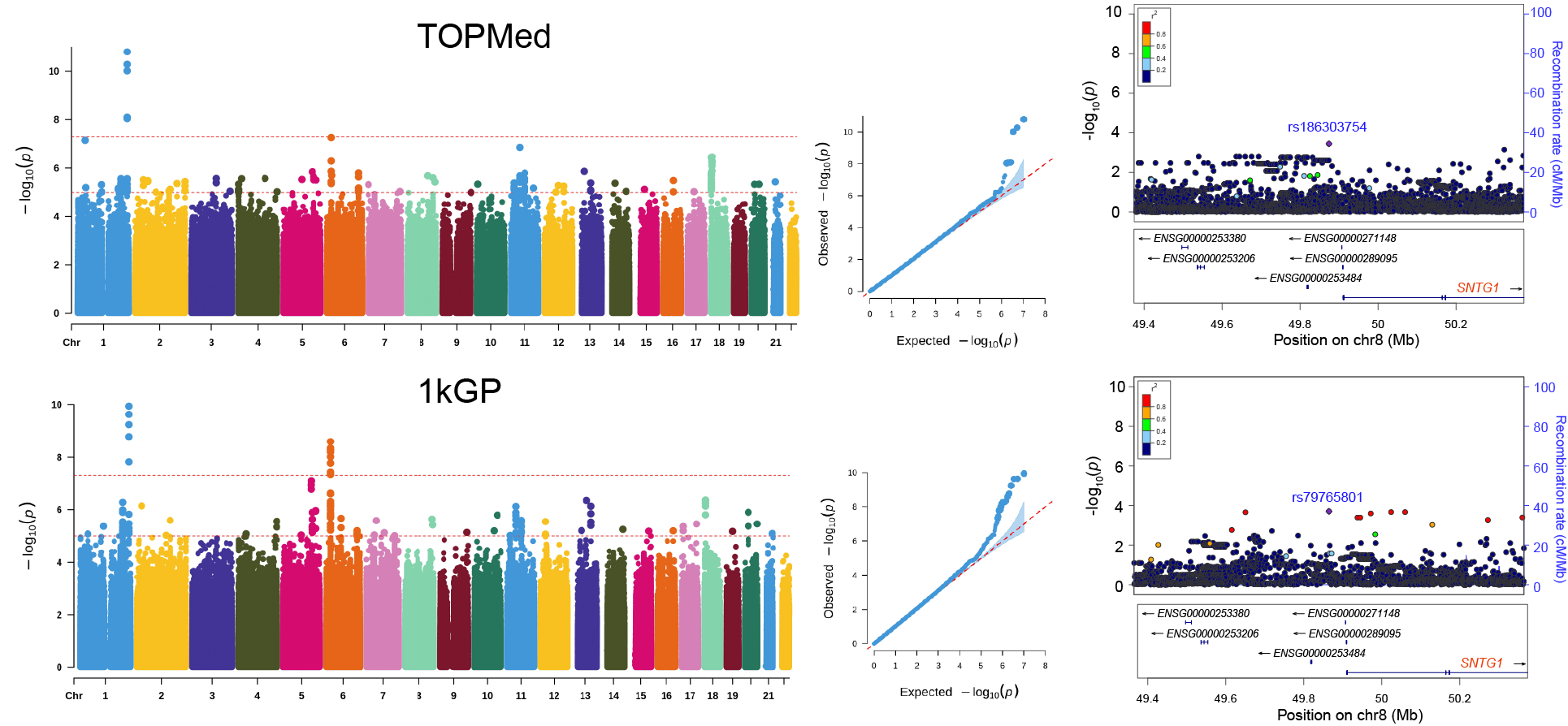
